# A Phase 2/3 observer-blind, randomized, controlled study to determine the safety and immunogenicity of SARS-CoV-2 recombinant spike protein vaccine in Indian children and adolescents aged 2 to 17 years

**DOI:** 10.1101/2023.01.03.23284130

**Authors:** Bhagwat Gunale, Dhananjay Kapse, Sonali Kar, Ashish Bavdekar, Sunil Kohli, Sanjay Lalwani, Sushant Meshram, Abhishek Raut, Praveen Kulkarni, Clarence Samuel, Renuka Munshi, Madhu Gupta, Joyce Plested, Shane Cloney-Clarke, MingZhu Zhu, Melinda Pryor, Stephanie Hamilton, Madhuri Thakar, Ashwini Shete, Abhijeet Dharmadhikari, Chetanraj Bhamare, Umesh Shaligram, Cyrus S. Poonawalla, Raburn M. Mallory, Gregory M Glenn, Prasad S. Kulkarni, the COVOVAX-Ped Study Group

## Abstract

**Background:** A recombinant, adjuvanted COVID-19 vaccine, SII-NVX-CoV2373 was manufactured in India and evaluated in Indian children and adolescents to assess safety and immunogenicity.

**Methods:** This was a Phase 2/3 observer-blind, randomized, controlled immuno-bridging study in children and adolescents aged 2 to 17 years. Participants were randomly assigned in 3:1 ratio to receive two doses of SII-NVX-CoV2373 or placebo on day 1 and day 22. Solicited adverse events (AEs) were collected for 7 days after each vaccination. Unsolicited AEs were collected for 35 days following first dose and serious AEs (SAEs) and adverse events of special interest (AESI) were collected throughout the study. Anti S IgG and neutralizing antibodies against the SARS-CoV-2 were measured at baseline, day 22, day 36 and day 180. Variant immune responses were assessed in a subset of participants at baseline, day 36 and day 180. Primary objectives were to demonstrate non-inferiority of SII-NVX-CoV2373 in each pediatric age group (12 to 17 years and 2 to 11 years, separately) to that in adults in terms of ratio of titers of both anti-S IgG and neutralizing antibodies 14 days after the second dose (day 36). Non-inferiority was to be concluded if the lower bound of 95% CI of the ratio was >0.67.

**Results:** A total of 920 children and adolescents (460 in each age cohort; 12 to 17 years and 2 to 11 years) were randomized and vaccinated. The demographic and baseline characteristics between the two groups were comparable in both age groups. After the second dose, there were more than 100-fold rise in anti-S IgG GMEUs and more than 84-fold rise in neutralizing antibodies GMTs from baseline in the participants who received SII-NVX-CoV2373. The lower bound of 95% CI of GMT ratios for anti-S IgG GMEUs and neutralizing antibodies in both age groups to those observed in Indian adults were >0.67, thus non-inferiority was met [Anti-S IgG GMT ratios 1.52 (1.38, 1.67), 1.20 (1.08, 1.34) and neutralizing antibodies GMT ratios 1.93 (1.70, 2.18), 1.33 (1.17, 1.50) for 2 to 11 years and 12 to 17 years groups, respectively]. The seroconversion rate was ≥ 98% (anti-S IgG) and ≥ 97.9 % (neutralizing antibodies) in both age groups, respectively. Similar findings were seen in the baseline seronegative participants. SII-NVX-CoV2373 also showed robust responses against various variants of concern. Injection site pain, tenderness, swelling, erythema and fever, headache, malaise, fatigue, were the common (≥5%) solicited adverse events which were transient and resolved without any sequelae. Throughout the study, only two causally unrelated SAEs and no AESI were reported.

**Conclusion:** SII-NVX-CoV2373 has been found safe and well tolerated in children and adolescents of 2 to 17 years. The vaccine was highly immunogenic and the immune response was non-inferior to that in adults.

**Registration - CTRI No. CTRI/2021/02/031554**

## Introduction

In the COVID-19 pandemic, adults were at high risk of severe disease and deaths. While children are generally considered as a lower risk population [1-3], there have been cases of severe COVID-19 and also deaths reported in children. Many of these children had some underlying medical conditions [4,5]. Moreover, children are also known to be carriers of the virus and can spread the virus among the adults [6] and therefore, it is important to protect children from the virus.

Considering all of these issues, pediatric vaccination has been considered very important. National Health Service, United Kingdom, US Centers for Disease Control and prevention and many other medical bodies recommend vaccination of the pediatric population [7,8]. Two mRNA vaccines have been approved in the USA for the age group of 6 months to 17 years [9]. In India, an inactivated vaccine and a receptor binding domain (RBD) based vaccines have also been approved for pediatric population from the age of 5 years and above [10], though the government program has included vaccination from 12 to 17 years [11]. As of 13 Jan 2023, around 42 million children of 12-14 years and 63 million children of 15-17 years have taken COVID-19 vaccines [12].

NVX-CoV2373, a SARS-CoV-2 recombinant spike protein nanoparticle vaccine was originally developed by Novavax in the USA. The vaccine has been found safe and has also shown high efficacy of 90% in adults [12,13] when the Wuhan strain was dominant. After a technology transfer, the vaccine is also manufactured in India by Serum Institute of India Pvt. Ltd. (SIIPL). This vaccine (SII-NVX-COV2373) has been tested in Indian adults and was found safe and immunogenic with more than 92% seroconversion with different antibody assays [14]. The vaccine has been granted emergency use authorization (EUA) for adults in India and received emergency use listing (EUL) by the World Health Organization in Dec 2021 [15,16]. SII-NVX-CoV2373 was subsequently evaluated in the pediatric population to immuno-bridge the vaccine with the adults. Based on this study, the vaccine was further approved for 12-17 years in March 2022 [17] and 7-11 years in June 2022 [18], respectively in India. The present paper summarizes the results of this study.

The primary objective of this study was to assess non-inferiority of SII-NVX-CoV2373 in adolescents of 12 to 17 years age and children of 2 to 11 years separately against that in the adult participants of the same study.^15^ Non-inferiority was to be demonstrated in terms of ratio of geometric mean of ELISA units (GMEU) of anti-spike IgG antibodies as well as geometric mean titres (GMTs) of neutralizing antibodies against SARS-CoV-2 at 14 days after the second dose. Another primary objective was to assess the safety of the study vaccine in comparison to placebo in terms of solicited local and systemic adverse events, unsolicited adverse events, serious adverse events and adverse events of special interest.

## Methods

The study was approved by the Indian regulatory authorities and Institutional Ethics Committees. The ICH Good Clinical Practice guidelines and the Declaration of Helsinki were followed. All children were enrolled after written informed consent of their parents, which was also audio visually recorded. In addition, a verbal assent from 7 to 11 years old children and a written assent for 12 to 17 years old children were also obtained.

This is a Phase 2/3 observer-blind, randomized, controlled study in children and adolescents of 2 to 17 years of age in India. A total of 920 eligible children and adolescents were enrolled across 10 tertiary care hospitals in this study, 460 in the 12 to 17 years age group and 460 in the 2 to 11 years age group. The participants were randomly assigned in 3:1 ratio to receive 2 doses of either SII-NVX-CoV2373 or placebo on day 1 and day 22. After vaccination, participants visited study sites on day 22 and day 36, day 85 and day 180. All participants were contacted telephonically on day 120 for safety follow up.

The recruitment happened in a stepwise manner. 100 participants were first enrolled in the 12 to 17 years age group. After review of the day 8 safety data by the Independent Data Safety Monitoring Board (DSMB) and the Indian regulatory authorities, the rest of the participants of this age group and 100 participants of the 7 to 11 years age group were enrolled. After the day 8 safety data of the first 100 participants from 7 to 11 years age group was reviewed by the DSMB and Indian regulatory authorities, the rest of the participants of this age group and 100 participants of the 2 to 6 years age group were enrolled. After day 8 safety data review by the DSMB and Indian regulatory authorities, remaining participants of the 2 to 6 years age group were enrolled. The recruitment of adults in Phase 3 study happened during March 2021 to July 2021, of 12 to 17 years was between Aug 2021 to Nov 2021; whereas of 2 to 11 years was between Sep 2021 to Feb 2022.

Safety was monitored during the study by onsite clinical staff. Safety data was reviewed at periodic intervals by the protocol safety review team (PSRT), an internal group of physicians, biostatisticians and a pharmacovigilance medical officer. In addition, an independent DSMB reviewed the safety data and provided an oversight on the study.

In all participants, approximately 6 ml blood was collected at baseline, day 22, day 36 and day 180 for immunogenicity testing. A nasopharyngeal swab was collected for detection of SARS-CoV-2 infection at baseline. Solicited, local and systemic adverse events (AEs) through 7 days after each dose were collected using diary cards. The parents were provided with digital thermometer, measuring scale and a diary.

### Study Population

Participants were healthy or medically stable children of ≥ 2 through 17 years of age. Female participants of 12 to 17 years of age who had attained menarche were required to have a negative urine pregnancy test at study entry. Children with any acute illness, history of COVID-19 disease, prior receipt of COVID-19 vaccine, severe allergic reactions, immunocompromised condition, were excluded from the study.

### Randomization and blinding

The randomization scheme was generated using SAS software version 9.4 (SAS Institute Inc, USA) for Interactive Response Technology (IRT) with block size of 6 and 3:1 allocation to SII-NVX-CoV2373 or placebo. The study participants, the study personnel responsible for the evaluation of any study endpoints and the laboratories involved in the immunogenicity testing were blinded to the treatment allocation. Personnel involved in getting randomization code by accessing interactive web response system (IWRS) and administration were unblinded and they did not conduct any study evaluations.

The unblinded monitors and statisticians from the CRO could access the participants level unblinded data as per the need.

After the vaccines were authorized for use and available through the program in the specific age group, the blind was broken on or after day 85 visit and participants were offered SII-NVX-CoV2373, if they had received Placebo and they were continued in the study only for safety follow-up.

### Study Products

The study vaccines were administered intramuscularly in the deltoid or anterolateral aspect of thigh as two doses of 0.5 ml each on day 1 and day 22 (+7 days). Covovax™ (SII-NVX-CoV2373 manufactured by SIIPL) is available as ready to use liquid formulation in a 10-dose vial. Each single dose of 0.5 ml contains 5 μg antigen and 50 μg Matrix-M**™** adjuvant (Batch No. 4301Z001, Manufacturing date July 2021, manufactured by SIIPL). A solution of 0.9% sodium chloride (NaCl) was used as a placebo (Batch No. 0700I002, Manufacturing date January 2020, manufactured by SIIPL).

### Safety assessment

The solicited AEs were collected for 7 days after each vaccination. The solicited local AEs included pain, tenderness, erythema, swelling and induration. The solicited systemic AEs included fever, headache, fatigue, malaise, arthralgia, myalgia, nausea and vomiting. Unsolicited adverse events were collected for 35 days after the first dose (14 days after second dose). Serious AEs (SAEs), related medically attended AEs (MAAEs) and AESI were collected throughout the 180 days of study participation. AESIs included potential immune-mediated conditions (PIMMCs), adverse events specific to COVID-19 or other potential adverse events that may have been determined at any time by regulatory authorities (Refer Protocol in Supplementary Appendix).

At any time during the study, if any participant presented with symptoms of suspected COVID-19 or had a history of contact with a confirmed COVID-19 case, the participant was tested by RT-PCR for SARS-CoV-2.

### Immunogenicity Assessment

Immunogenicity was assessed by Anti-Spike (anti-S) IgG antibodies and neutralizing antibodies against SARS-CoV-2 to prototype strain on days 1, 22, 36 and 180. Anti-S IgG antibodies were measured by validated ELISA assay (Novavax Clinical Immunology, Gaithersburg, USA) in all participants.

Neutralizing antibodies against SARS-CoV-2 were measured using a validated virus neutralizing assay (VNA) with wild type virus (SARS-CoV-2 hCoV-19/Australia/ VIC01/2020, GenBank MT007544.1) on day 1 and day 36 in all participants and in a randomly selected subset of participants (maintaining 3:1 allocation) on day 22 and day 180 (213 samples and 132 samples for day 22 in 12 to 17 years and 2 to 11 years groups, respectively; 137 samples and 120 samples for day 180 in 12 to 17 years and 2 to 11 years groups, respectively) as exploratory objectives. In addition, neutralizing antibodies against omicron BA.1 (B.1.1.529) were measured using a validated VNA in randomly selected subset of 75 participants from 12 to 17 years group and 80 participants from 2 to 11 years group (maintaining 3:1 allocation) (360Biolabs, Melbourne, Australia).

Anti-N (nucleocapsid) IgG antibodies were measured at baseline, days 22, 85 and 180 in all participants by Chemiluminescence Microparticle immune assay (CMIA) using ARCHITECT SARS-CoV-2 IgG kit (6R86) and ARCHITECT i2000 SR system from Abbott Laboratories (NARI-Indian Council of Medical Research, Pune, India) using ARCHITECT SARS-CoV-2 IgG kit (6R86) (Abbott). The cutoff for seropositivity was S/C index ≥ 1.40.

Immunogenicity against Delta, Omicron BA.1 and Omicron BA.5 variants were also assessed in a randomly selected subset of 44 participants from 12 to 17 years group and 53 participants from 2 to 11 years group (maintaining 3:1 allocation) by anti-S IgG antibodies by ELISA assay on days 1, 36 and 180. In addition, immune response against ancestral strain (Wuhan), Delta, Omicron BA.1 and Omicron BA.5 variants were determined in a subset by human ACE2 (hACE2) receptor binding inhibition assay on days 1, 36 and 180 in the same subset. Both assays were conducted at Novavax Clinical Immunology, Gaithersburg, USA.

### Statistical Analysis

The study had 82% power to detect at least one serious adverse event among 690 children and adolescents administered SII-NVX-CoV2373, if the frequency of causally related SAEs was 1 in 400. In each of the pediatric cohort age group (2 to 11 years and 12 to 17 years), 345 participants received the SII-NVX-CoV2373 vaccine and 115 received placebo. The proportion of non-evaluable participants was assumed to be ≤ 20%, which led to sample size 276 evaluable participants. The study had over 99% statistical power to achieve a non-inferiority objective for 2-11 years and 12-18 years groups separately in terms of GMEU ratio compared with 276 adult participants who had received SII-NVX-CoV2373. Non-inferiority was to be concluded if the lower limit of the two-sided 95% CI for the GMEU ratio for Anti-S IgG antibodies and GMT) ratio of neutralizing antibodies for SII-NVX-CoV2373 between each pediatric age group (2 to 11 years) and (12 to 17 years) and adult cohort was more than 0.67, as per the WHO guidelines for clinical evaluation of vaccines [19]. Additional assumptions were one sided significance level of 0.025, a zero difference in the antibody titers between pediatric and adult participants (i.e. a GMEU / GMT ratio of 1 and coefficient variation of 1.35. Sample size calculations were performed using a non-inferiority test for the ratio of two means in PASS 15.0.7 Version software. Multiple Imputation model with classification variables vaccine, sex, and continuous covariates log baseline titer of anti-S IgG and neutralizing antibodies were used to impute 50 values for each missing value when non-inferiority was analysed. All participants with positive RT PCR for SARS CoV 2 at baseline or at any point through study were excluded from immunogenicity analysis population and were part of full analysis population.

All participants who received at least one dose of study vaccine were part of the safety population. The Immunogenicity population consisted of all participants who received the first dose of the study vaccine and provided an evaluable serum sample for at least one assessment and had baseline data available. The immunogenicity population excluded any data from time points following of SARS-CoV-2 infection or from participants with major protocol deviations impacting the study outcomes. Those participants for whom at least one post-vaccination immunogenicity data was not available, there were excluded from immunogenicity analysis population. All the participants from the placebo group who were unblinded on or after day 85 visit for COVID-19 vaccination were censored from the group-wise data analysis and their data was analyzed separately.

For the primary immunogenicity analysis, ANCOVA fitted to the log transformed anti-S IgG as well as neutralizing antibodies with terms for vaccine group, log baseline titer and sex to compare each pediatric cohort versus adult cohort participants vaccinated with SII-NVX-CoV2373 for primary endpoint.

Categorical data are expressed as proportions while quantitative data are expressed as mean and SD.

All statistical analyses were performed using SAS® Software version 9.4 or later.

## RESULTS

This analysis includes data collected for the entire study i.e. until the day 180 visit of all participants from ≥ 12 to < 18 (12 to 17) years of age and ≥ 2 to < 12 (2 to 11) years of age.

### Disposition of Participants

A total of 470 participants 12 to 17 years of age were enrolled (screened) and 461 were randomized. Among these 461 participants, 460 received the first dose of study vaccine (346 SII-NVX-CoV2373 and 114 Placebo) and 445 received the second dose of study vaccine (335 SII-NVX-CoV2373 and 110 Placebo). Fifteen participants withdrew the consent before the second dose. The safety population comprised all 460 participants and immunogenicity analysis population comprised 441 participants (Figure 1). The demographic and baseline characteristics of the two groups are given in Table 1.

**Figure 1:**
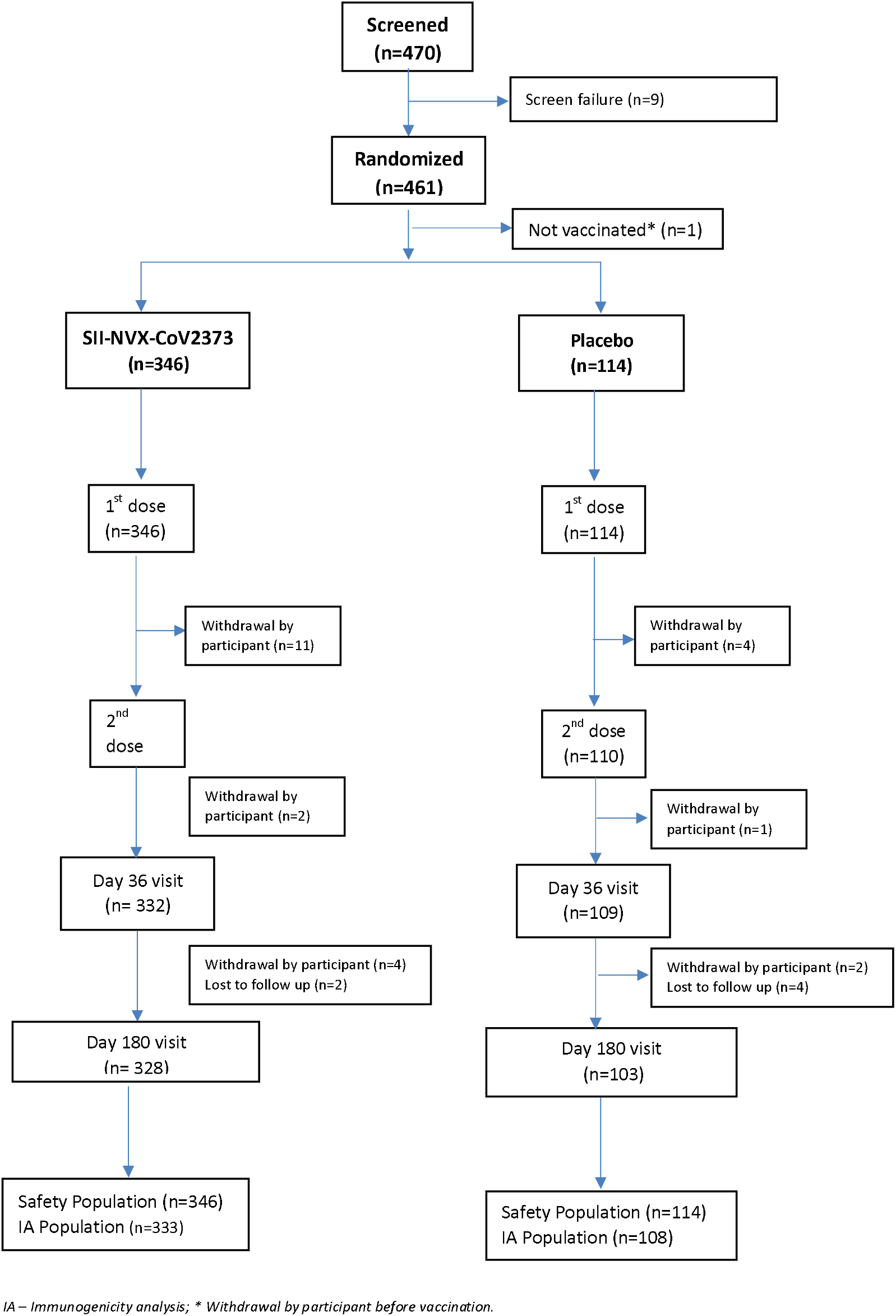
CONSORT Flow Chart – 12 to 17 years.

**Table 1:** Demographics and Baseline Characteristics – (Safety Population)

| Parameter | 2-11 years |  | 12-17 years |  |
| --- | --- | --- | --- | --- |
|  | SII-NVX-CoV2373<br>(N=345) | Placebo<br>(N=115) | SII-NVX-CoV2373<br>(N=346) | Placebo<br>(N=114) |
| Age (Years), Mean (SD) | 6.7 (2.66) | 6.5 (2.70) | 14.3 (1.55) | 14.3 (1.64) |
| Sex, n (%) |  |  |  |  |
| Male | 176 (51.0) | 53 (46.1) | 188 (54.3) | 54 (47.4) |
| Female | 169 (49.0) | 62 (53.9) | 158 (45.7) | 60 (52.6) |
| BMI (SD) | 15.64 (2.934) | 15.61 (3.040) | 19.47 (4.481) | 19.95 (4.833) |
| Baseline serology and/or Positive<br>RT-PCR results for SARS-CoV-2, n (%)* | 37 (10.7) | 16 (13.9) | 45 (13.0) | 14 (12.3) |
| Negative | 306 (88.7) | 98 (85.2) | 300 (86.7) | 100 (87.7) |

\*Results missing for 3 participants (2 to 11 years) and 1 participant (12 to 17 years), respectively.
SD – Standard Deviation

A total of 474 participants 2 to 11 years of age were enrolled (screened) and 460 were randomized and received the first dose of study vaccine (345 SII-NVX-CoV2373 and 115 Placebo) and 445 received the second dose of study vaccine (333 SII-NVX-CoV2373 and 112 Placebo). Fifteen participants withdrew before the second dose. The safety population comprised all 460 participants and immunogenicity analysis population comprised 437 participants (Figure 2). The demographic and baseline characteristics between the two groups were comparable [Table 1].

**Figure 2:**
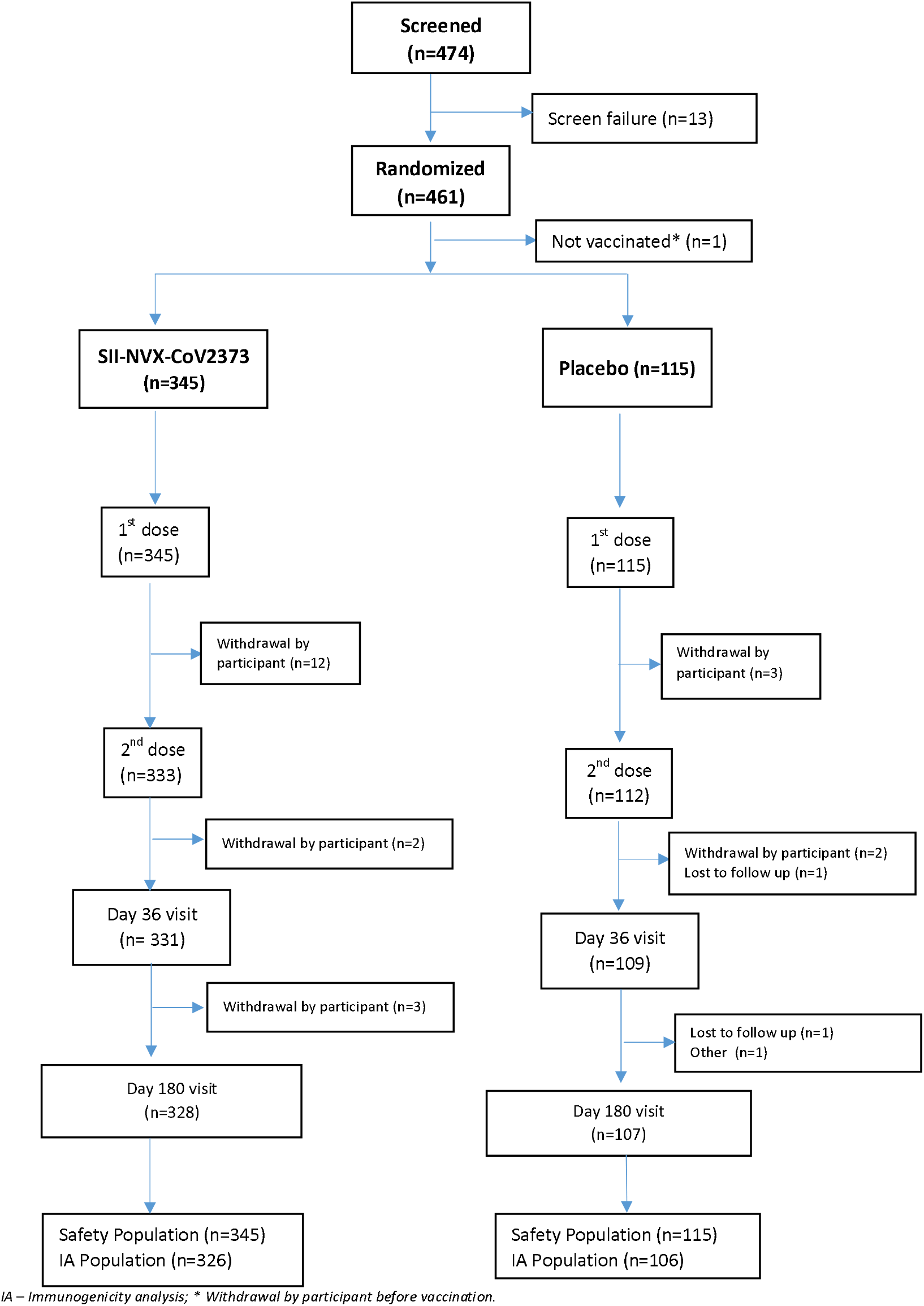
CONSORT Flow Chart – 2 to 11 years.

### Immunogenicity Results

#### Anti-S IgG against ancestral strain (Wuhan)

Baseline GMEUs of anti-S IgG to prototype strain were comparable between the SII-NVX-CoV2373 and the Placebo groups in both 12-17 years and 2-11 years age group. After the second dose, there was 101.9 fold-rise (12-17 years) and 169.2 fold-rise (2-11 years) in the anti-S IgG in the SII-NVX-CoV2373 group. There was negligible rise in the placebo groups. On day 180, there was a decline in titres, but still they were 37-fold higher (12-17 years) and 31-fold higher (2-11 years) than the baseline in the SII-NVX-CoV2373 group. The corresponding values in the placebo group were 8.1 fold (12-17 years) and 3.4-fold (2-11 years) [Table 3]. The fold-rises in the baseline negative participants (negative for both anti-N IgG and RT-PCR for SARS CoV-2), were more than 123-fold in both the age groups. On day 180, they were still more than 38-fold higher than the baseline in the SII-NVX-CoV2373 group in both the age groups. The corresponding values in the placebo group was more than 4-fold in both the age groups [Table S1].

Non-inferiority for SII-NVX-CoV2373 for each pediatric cohort separately (12-17 years and 2-11 years) in comparison to the adult cohort was met using anti-S IgG titers, with 1.08 as the lower bound of 95% CI in the 12 to 17 years age group and with 1.38 as the lower bound of 95% CI in the 2-11 year age group [Table 2].

**Table 2:** Non-inferiority of COVOVAX™ (Pediatric) to COVOVAX™ (Adult) in terms of Anti-S IgG and neutralizing antibodies at Visit 3-Day 36 (Immunogenicity Analysis Population)

|  | 2 to 11 years |  | 12-17 years |  |
| --- | --- | --- | --- | --- |
| Multiple Imputation [1] Results<br>Using Rubin’s Method [2] | COVOVAX<sup>PD</sup><br>(N=326) | COVOVAX<sup>AD</sup><br>(N=340) | COVOVAX<sup>PD</sup><br>(N=333) | COVOVAX<sup>AD</sup><br>(N=340) |
| Anti-S IgG |  |  |  |  |
| GMEU [3] with 95% CI | 216369.87<br>(202056.83, 231696.80) | 142586.82<br>(133280.59, 152542.85) | 171204.19<br>(158598.33, 184812.00) | 142122.16<br>(131793.34, 153260.47) |
| GMEU Ratio [3] with 95% CI | 1.52 (1.38, 1.67) |  | 1.20 (1.08, 1.34) |  |
| Neutralizing antibodies |  |  |  |  |
| GMT [3] with 95% CI | 6940.82<br>(6349.74, 7586.93) | 3602.39<br>(3300.10, 3932.37) | 4732.36<br>(4323.63, 5179.73) | 3568.04<br>(3264.78, 3899.48) |
| GMT Ratio [3] with 95% CI | 1.93 (1.70, 2.18) |  | 1.33 (1.17, 1.50) |  |

[1] Multiple Imputation model with classification variables vaccine, sex, and continuous covariates log baseline titer used to impute 50 values for each missing value.
[2] Rubin's method in PROC MIANALYZE used to pool estimates and SE across the 50 multiply imputed datasets.
[3] Pooled ANCOVA results, LS Mean and its 95% CI values by treatment were used to generate the GMEU / GMT and 95% CI and the differences in LS Means and corresponding 95% CI limits were used to obtain GMEU / GMT Ratio and 95% CI using back transforming to the original scale.
PD – Pediatric cohort, AD – Adult cohort. Specified age group detail is applicable for pediatric cohort (i.e. 12 to 17 years and 2 to 11 years).

**Table 3:**
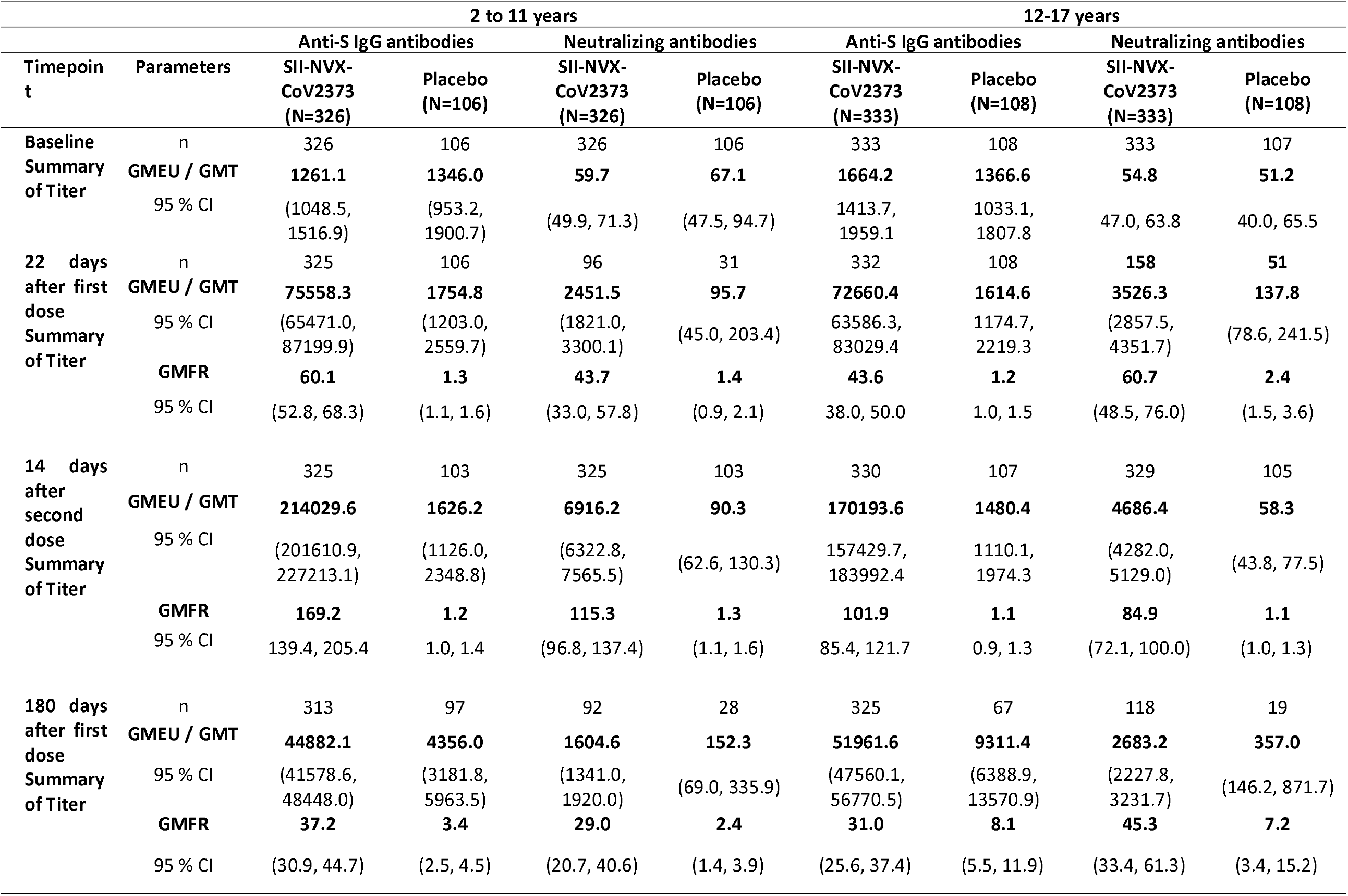
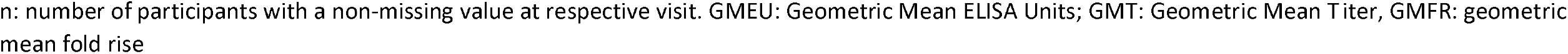
Summary of Anti-S IgG antibodies and Neutralizing antibodies (nAb) against SARS-CoV-2 (Immunogenicity analysis population)

On day 36, there was 98.8% seroconversion (12-17 years) and 99.1% (2-11 years) after the second dose in the SII-NVX-CoV2373 group and it was less than 7.8% in the Placebo group. On day 180, the seroconversion was still more than 90% in both the age groups who received SII-NVX-CoV2373. In the placebo group, this increased to 43% and 73% in 2-11 year and 12-17 year age groups, respectively [Table 4]. In the baseline seronegative participants on day 36, this was 99.3% (12-17 years) and 100% (2-11 years) in the SII-NVX-CoV2373 group while it was less than 8.7% in the Placebo group. In this population too, the seroconversion rate was still more than 90% in both the age groups on day 180. In the placebo group, this increased to 43% and 73% in 2-11 year and 12-17-year age groups, respectively [Table S2].

**Table 4:**
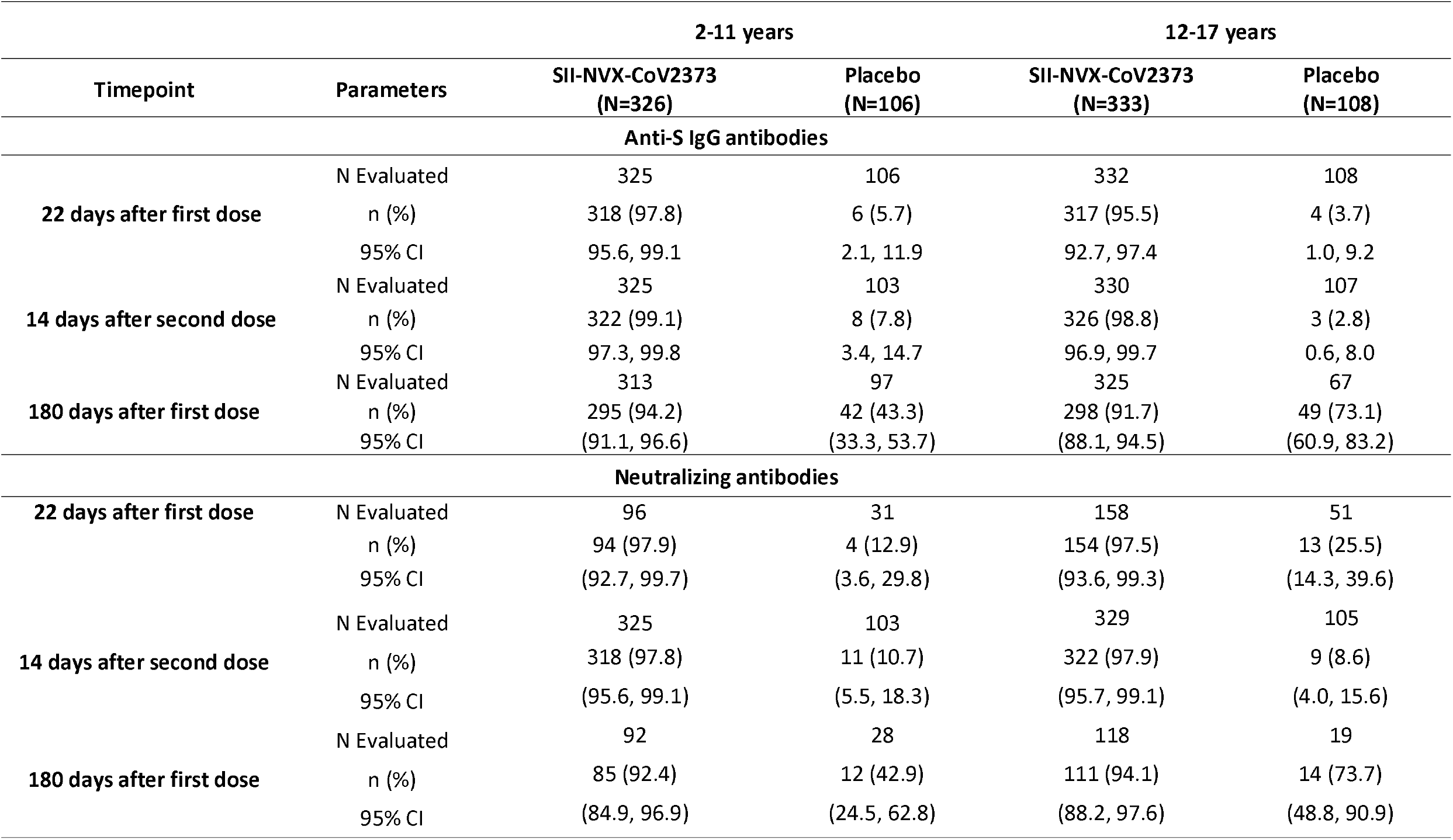
Proportion of Participants with Seroconversion for Anti-S IgG Antibodies and neutralizing antibodies (Immunogenicity analysis population)

### Neutralizing antibodies (nAbs) against ancestral strain (Wuhan)

Baseline GMTs of neutralizing antibodies (nAbs) were comparable between the SII-NVX-CoV2373 and the Placebo group in both the age groups. After the second dose, there was 84.9 fold-rise (12-17 years) and 115.3 fold-rise (2-11 years) in nAbs GMTs from the baseline in the SII-NVX-CoV2373 group. On day 180, there was still 45.3-fold-rise (12-17 years) and 29-fold-rise (2-11 years) in the SII-NVX-CoV2373 group [Table 3]. In the baseline seronegative participants, the corresponding values were more than 98-fold (12-17 years) and 138-fold (2-11 years) on day 36 and more than 54-fold (12-17 years) and 34-fold (2-11 years) on day 180. There was no rise in the Placebo group on day 36, whereas 7.4-fold rise (12-17 years) and 2.9-fold rise (2-11 years) from the baseline was seen on day 180 [Table S1].

Non-inferiority for each pediatric cohort in comparison to the adult cohort was met, with 1.17 and 1.70 as the lower bound of 95% CI of GMT ratio in 12-17 years and 2-11 years age groups, respectively [Table 2].

On day 36, there was more than 98% seroconversion after the second dose in the SII-NVX-CoV2373 group in both age groups while it was more than 91% on day 180. [Table 4]. It was more than 98% and more than 94% in baseline negative participants in both age groups, on days 36 and 180, respectively. In the same population, there was less than 12% seroconversion in the placebo group on day 36 and 76.5% in 12-17 years and 41.7% in 2-11 years age groups on day 180 [Table S2].

### Immunogenicity against variants

Compared to baseline, there was around 70- to 100-fold rise and 17- to 40-fold rise in anti-S IgG antibodies against Delta, Omicron BA.1 and Omicron BA.5 variants on day 36 and day 180, respectively in the SII-NVX-CoV2373group [Table S3].

In terms of hACE2 receptor binding inhibition GMTs, there was around 20- to 40-fold rise and 9-fold rise against Delta, Omicron BA.1 and Omicron BA.5 variants on day 36 and day 180, respectively in the SII-NVX-CoV2373 group. For the Wuhan strain, the rise was 67- and 13.2-fold on day 36 and day 180, respectively [Table S4].

#### COVID-19 incidence

At 14 days after the first dose, there were 3 cases of symptomatic COVID-19 (0.9%) in the SII-NVX-CoV2373 group and 2 (1.8%) cases of symptomatic COVID-19 and one case of asymptomatic SARS CoV-2 infection (0.9%) in the placebo group in the 12 to 17 year group, and there was 1 each case of symptomatic COVID-19 in the SII-NVX-CoV2373 group (0.3%) and placebo group (0.9%) in the 2 to 11 year group. From 14 days after the second dose until day 180 visit, there were 2 cases of symptomatic COVID-19 (0.6%) and 2 cases of asymptomatic SARS CoV-2 infection (0.6%) in the SII-NVX-CoV2373 group and 2 (1.8%) cases of symptomatic COVID-19 and one case of asymptomatic SARS CoV-2 infection (0.9%) in the placebo group in the 12 to 17 year group, whereas there was 1 each case of symptomatic COVID-19 in the SII-NVX-CoV2373 group (0.3%) and placebo group (0.9%) in the 2 to 11 year group. No case was severe, none required ICU admission and all recovered completely.

### Safety Results

#### SAEs and AESIs

Among 12-17 year adolescents, two SAEs were reported in vaccine recipients which were viral infection and gastroenteritis. Both these events were assessed as causally unrelated to the study vaccine. No AESI was reported in the study. Among 2 to 11 year old children, there were no SAEs or AESIs reported.

##### Unsolicited AEs

In the 12 to 17 year age group, a total of 30 AEs were reported in 28 participants (8.1%) in the SII-NVX-CoV2373 group and 12 AEs in 6 participants (5.3%) in the Placebo group. None of these were considered causally related to the study vaccines except for the event of diarrhea in the Placebo group.

In the 2 to 11 year age group, a total of 49 AEs were reported in 39 participants (11.3%) in the SII-NVX-CoV2373 group and 14 AEs in 13 participants (11.3%) in the Placebo group. None of these were considered causally related to the study vaccines except for the three events (1 each of pyrexia, allergic dermatitis, and diarrhea) in the SII-NVX-CoV2373 group and one in the placebo group (diarrhea).

Almost all AEs were of mild severity and all resolved without any sequelae.

##### Solicited AEs

In the 12 to 17 year age group, after the first dose, there were 271 solicited AEs reported in 138 participants (39.9%) in the SII-NVX-CoV2373 group whereas 54 solicited AEs were reported in 26 participants (22.8%) in the placebo group. After the second dose, there were 322 solicited AEs reported in 126 participants (37.6%) in the SII-NVX-CoV2373 group whereas 41 solicited AEs were reported in 19 participants (17.3%) in the placebo group. The common AEs in both the groups included injection site pain, tenderness, headache, fatigue, fever [Table 5].

**Table 5:** Summary of Solicited AEs (Safety Population)

|  | 2-11 years |  |  |  | 12-17 years |  |  |  |
| --- | --- | --- | --- | --- | --- | --- | --- | --- |
|  | First dose |  | Second dose |  | First dose |  | Second dose |  |
|  | SII-NVX-CoV2373<br>(N=345) | Placebo<br>(N=115) | SII-NVX-CoV2373<br>(N=333) | Placebo<br>(N=112) | SII-NVX-CoV2373<br>(N=346) | Placebo<br>(N=114) | SII-NVX-CoV2373<br>(N=335) | Placebo<br>(N=110) |
|  | n (%) [E] | n (%) [E] | n (%) [E] | n (%) [E] | n (%) [E] | n (%) [E] | n (%) [E] | n (%) [E] |
| Participants with at Least One Solicited AE | 122 (35.4) [248] | 15 (13.0) [29] | 163 (48.9) [367] | 13 (11.6) [27] | 139 (40.2) [272] | 26 (22.8) [54] | 129 (38.5) [329] | 20 (18.2) [42] |
| Participants with at Least One Local Solicited AE | 84 (24.3) [118] | 10 ( 8.7) [15] | 87 (26.1) [136] | 9 ( 8.0) [17] | 90 (26.0) [123] | 12 (10.5) [16] | 86 (25.7) [140] | 10 ( 9.1) [13] |
| Injection Site Pain | 81 (23.5) [81] | 9 ( 7.8) [9] | 83 (24.9) [83] | 9 ( 8.0) [9] | 85 (24.6) [85] | 11 ( 9.6) [11] | 82 (24.5) [82] | 9 (8.2) [9] |
| Injection Site Tenderness | 27 ( 7.8) [27] | 3 ( 2.6) [3] | 20 ( 6.0) [20] | 1 ( 0.9) [1] | 23 ( 6.6) [23] | 1 ( 0.9) [1] | 29 ( 8.7) [29] | 3 (2.7) [3] |
| Injection Site Swelling | 6 ( 1.7) [6] | 2 ( 1.7) [2] | 14 ( 4.2) [14] | 2 ( 1.8) [2] | 8 ( 2.3) [8] | 2 ( 1.8) [2] | 13 (3.9) [13] | 0 |
| Injection Site Induration | 2 ( 0.6) [2] | 1 ( 0.9) [1] | 5 ( 1.5) [5] | 2 ( 1.8) [2] | 7 (2.0) [7] | 1 ( 0.9) [1] | 10 (3.0) [10] | 1 (0.9) [1] |
| Injection Site Erythema | 2 ( 0.6) [2] | 0 | 14 ( 4.2) [14] | 3 ( 2.7) [3] | 0 | 1 ( 0.9) [1] | 6 (1.8) [6] | 0 |
| Participants with at Least One Systemic Solicited AE | 80 (23.2) [130] | 10 ( 8.7) [14] | 133 (39.9) [231] | 8 ( 7.1) [10] | 87 (25.1) [149] | 19 (16.7) [38] | 99 (29.6) [189] | 17 (15.5) [29] |
| Fever | 50 (14.5) [50] | 6 ( 5.2) [6] | 99 (29.7) [99] | 1 ( 0.9) [1] | 27 (7.8) [27] | 7 (6.1) [7] | 58 (17.3) [58] | 3 (2.7) [3] |
| Headache | 19 ( 5.5) [19] | 2 ( 1.7) [2] | 37 (11.1) [37] | 4 ( 3.6) [4] | 39 (11.3) [39] | 11 ( 9.6) [11] | 42 (12.5) [42] | 9 (8.2) [9] |
| Malaise | 17 ( 4.9) [17] | 2 ( 1.7) [2] | 19 ( 5.7) [19] | 1 ( 0.9) [1] | 14 (4.0) [14] | 2 (1.8) [2] | 23 ( 6.9) [23] | 2 (1.8) [2] |
| Fatigue | 15 ( 4.3) [15] | 1 ( 0.9) [1] | 18 ( 5.4) [18] | 2 ( 1.8) [2] | 30 (8.7) [30] | 5 (4.4) [5] | 30 ( 9.0) [30] | 4 (3.6) [4] |
| Myalgia | 12 ( 3.5) [12] | 2 ( 1.7) [2] | 14 ( 4.2) [14] | 1 ( 0.9) [1] | 16 (4.6) [16] | 5 (4.4) [5] | 14 (4.2) [14] | 4 (3.6) [4] |
| Arthralgia | 4 ( 1.2) [4] | 0 | 11 ( 3.3) [11] | 0 | 15 (4.3) [15] | 3 (2.6) [3] | 10 (3.0) [10] | 4 (3.6) [4] |
| Nausea | 5 ( 1.4) [5] | 0 | 20 ( 6.0) [20] | 0 | 7 (2.0) [7] | 2 (1.8) [2] | 8 (2.4) [8] | 2 (1.8) [2] |
| Vomiting | 8 ( 2.3) [8] | 1 ( 0.9) [1] | 13 ( 3.9) [13] | 1 ( 0.9) [1] | 1 (0.3) [1] | 3 (2.6) [3] | 4 (1.2) [4] | 1 (0.9) [1] |

In the 2 to 11 year age group, there were 248 solicited AEs reported in 122 participants (35.4%) in the SII-NVX-CoV2373 group whereas 29 solicited AEs were reported in 15 participants (13.0%) in the Placebo group after the first dose. There were 367 solicited AEs reported in 163 participants (48.9%) in the SII-NVX-CoV2373 group whereas 27 solicited AEs were reported in 13 participants (11.6%) in the Placebo group after the second dose. In this age group as well, the common AEs in both the groups included injection site pain, tenderness, headache, fatigue, fever [Table 5].

Almost all AEs were of mild severity and all resolved without any sequalae. Most of the solicited AEs started within 1 day and lasted for 1 to 2 days in both the age groups.

## Discussion

The present study evaluated safety and immunogenicity of SII-NVX-CoV2373 in Indian children and adolescents of 2-17 years. The vaccine was highly immunogenic with more than 98% seroconversion for both anti-S IgG and neutralizing antibodies in adolescents of 12 to 17 years of age and children of 2 to 11 years of age. There was a more than 84-fold rise from baseline for both anti-S IgG and neutralizing antibodies. No AESI was reported and no causally related SAE was reported. SII-NVX-CoV2373 is safe and well tolerated in the pediatric population of 2-17 years of age.

An age effect was clearly seen in the immune responses. Not only did both 12 to 17 years adolescents and 2 to 11 years children age groups show non-inferior immune responses to adults, but in fact, the titers in both the pediatric age groups were higher than in adults [14]. Moreover, the titers in the 2 to 11 years age group were also higher than in the 12 to 17 years age group. It is known that the younger populations mount higher immune response as compared to older population [20].

We demonstrated non-inferiority of SII-NVX-CoV2373 in children against the adult age group. We did this because at the time of the study, no vaccine was approved in India for pediatric age group. As a result, we had to use placebo as a control. The vaccine efficacy was already demonstrated in adults and therefore, we used adult results as a control so as to immuno-bridge the vaccine in children. This is the approach used in other COVID-19 vaccine studies [21-24].

To our knowledge this is the first study in India in which any recombinant protein COVID-19 vaccine has been tested between 2 to 5 years of age. mRNA vaccines have been tested and recommended for use in 6 month to 5 years population [8]. In general, there has been a concern of myocarditis with mRNA vaccines in younger populations [25-28]. So far SII-NVX-CoV2373 has not been found to cause any related SAE or AESI in this study as well as in previous studies conducted in India, though the size of the study populations has been small.

The day 180 results showed decline in IgG as well as neutralizing antibodies in the vaccine group, though they were still much higher than the baseline titres and much higher than the placebo group. Seroconversion was still more than 90%. There was a substantial rise in the titres in the placebo groups compared to baseline likely due to SARS-CoV-2 infections. These results are on expected lines as seen with SII-NVX-CoV2373 in adults [14].

The GMTs and seroconversion in placebo groups on day 180 were significantly higher as compared to baseline as well as day 36. This was probably due to the Omicron wave that affected all age groups in the country during December 2021-March 2022. Even there, the titres were higher in the 12-17 years age group as compared to 2-11 years. Again, this could be due to higher infections in older children as compared to younger children. None of these children received a COVID-19 vaccine nor reported a symptomatic COVID-19. As of 24 July 2022, children of 5-14 years and those less than 5 years of age had 10.44% and 2.47% infections during Omicron wave, respectively [29].

We also tested a subset of participants for immune response against variants of concern. SII-NVX-CoV2373 showed high GMTs as well as seroconversions for Delta, BA.1 and BA.5 variants though slightly lower than those for the Wuhan strain. This is known with other vaccines [30]. Still such high titres indicate that vaccines based on the Wuhan strain may give protection from various variants, especially for severe disease and deaths.

Pediatric COVID-19 vaccination has faced some degree of hesitancy among parents [31-33]. One study in Canada found that despite parents’ high COVID-19 vaccination uptake for themselves (88.8%), intentions for vaccinating children aged 5–11 years was low (56%) [34]. As a result, the vaccine coverage has not been optimum [35]. Some important reasons for the low coverage have been concerns about vaccine safety and effectiveness [36]. There is also a perception that the risk for serious COVID-19–associated illness is low in children [36]. On this background, the safety profile in children can be an advantage of the SII-NVX-CoV2373.

Baseline serology and/or RT-PCR positivity for SARS-CoV-2 was around 10-13% in both the pediatric age groups. The corresponding value in the Phase 3 study in adults was around 33% [14]. This probably means that the first wave in 2020 as well as the second wave in 2021 and third wave in 2021-2022 in India did not affect children significantly. However, another study conducted during March-June 2021 showed a seroprevalence of 55.7% in the <18 years age group and 63.5% in the ≥18 years age group in India [37]. While this study measured total antibodies against S-RBD SARS-CoV-2 virus, our study measured anti-N IgG antibodies. Disparities in prevalence estimate obtained using different serological assays are known [38].

NVX-CoV2373 has shown around 90% efficacy in adults [12,13]. The immune responses by SII-NVX-CoV2373 in children were much higher than that seen in adults [14]. Higher levels of all immune markers are known to correlate with a reduced risk of symptomatic infection [39]. Human challenge studies of seasonal coronaviruses reported high levels of baseline neutralizing antibodies in uninfected or asymptomatic people [40], thus indicating that high antibody titres provide protection from disease. Considering these factors, SII-NVX-CoV2373 is expected to give high degree of protection in children, at least as much as in adults.

Our study had a few potential limitations. We excluded previous known SARS-CoV-2 infection cases from the study. However, because of this, we could assess the actual vaccine effect and no response was seen in the placebo group until 14 days after the second dose. We did not assess efficacy of the vaccine in our study population. However, NVX-CoV2373 has already demonstrated efficacy in two large studies [12,13] and therefore, SII-NVX-Cov2373 with identical composition as NVX-Cov2373 could be bridged immunologically. Severe COVID-19 has been reported in children with some underlying medical conditions [5,6] but we excluded such children from our study. Immunogenicity of Covid-19 vaccines among such children and adolescents has been lower than healthy individuals [41-44] which is expected. However, it was still an acceptable immune response [44]. Recruitment in the study was scalonated, which probably meant that adults and adolescents/children were exposed to different strains, including variants of concerns during the follow up period. This may have affected the immunogenicity results.

To our knowledge there are only 6 published Phase 2/3 studies of authorized COVID-19 vaccines in pediatric population for primary immunization – 2 mRNA vaccines [21-24], 1 receptor binding domain vaccine [45] and 1 nonrandomized study of an inactivated vaccine [46]. Our study is the first published data in pediatric age group for the spike protein vaccine and adds to the existing evidence that COVID-19 vaccines work well in pediatric population.

To conclude, SII-NVX-COV2373 was safe and well tolerated in children of 2 to 17 years age group; the vaccine was highly immunogenic and showed higher immune response than in adults and also showed robust responses against various variants of concern. The vaccine may be used in the pediatric vaccination drive against COVID-19.

## Data Availability

All data produced in the present work are contained in the manuscript.

## Acknowledgements

We gratefully acknowledge the contribution by our DSMB members Drs. Sharad Agarkhedkar, Charudatta Joglekar and Ommen John. We acknowledge the study participants and their parents. We also acknowledge PPD for contribution to study monitoring, data management and statistical analysis for the study.

## Supplementary tables

**Table S1:**
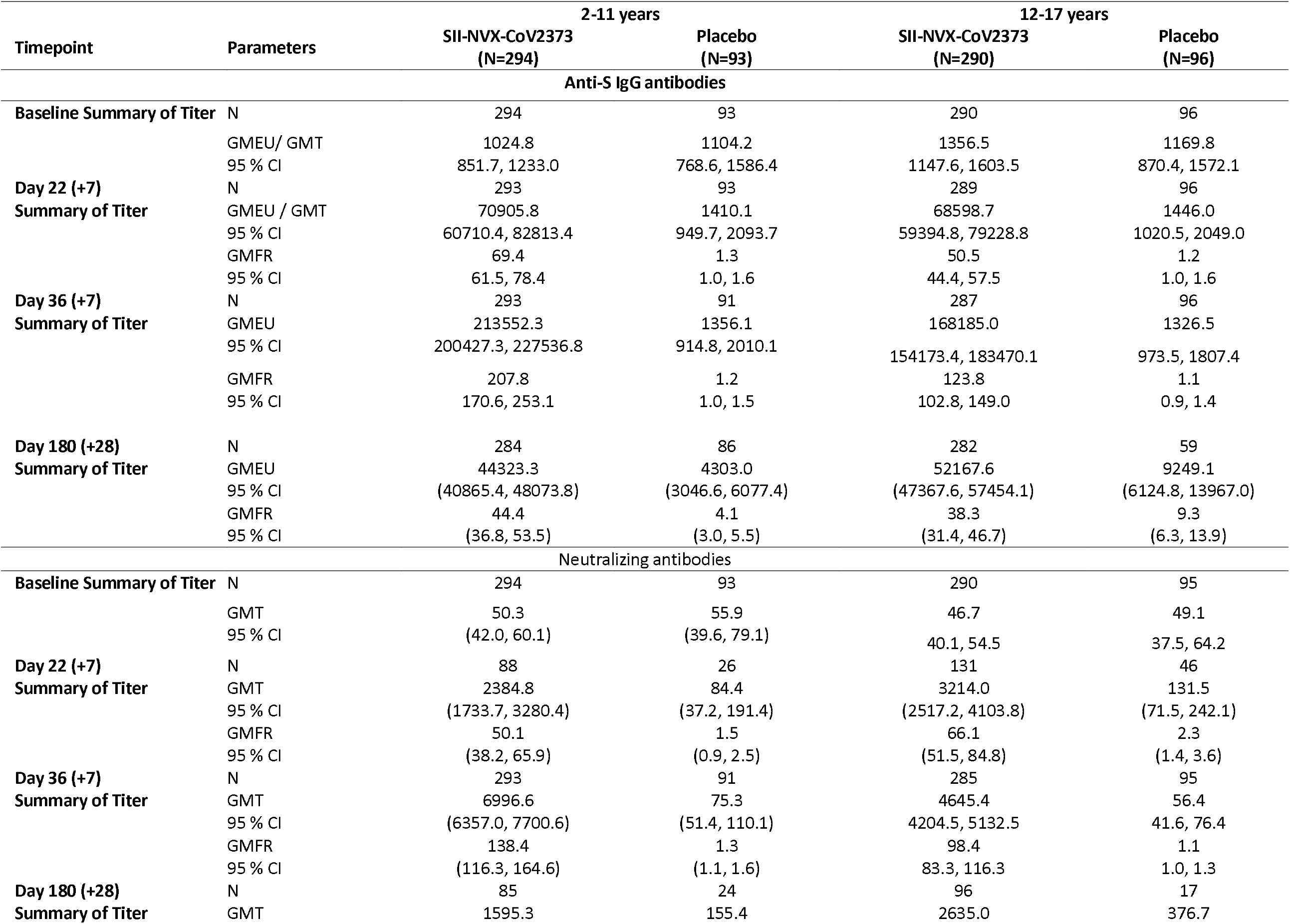
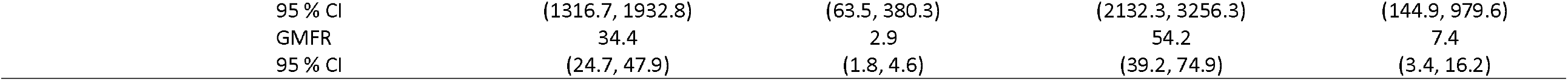
Summary of Anti-S IgG and neutralizing antibodies in baseline seronegative and RT-PCR negative (Immunogenicity Analysis Population)

**Table S2:** Proportion of Participants with Seroconversion for Anti-S IgG Antibodies and neutralizing antibodies in baseline seronegative and RT-PCR negative (Immunogenicity Analysis Population)

| Timepoint | Parameters | 2-11 years |  | 12-17 years |  |
| --- | --- | --- | --- | --- | --- |
|  |  | SII-NVX-CoV2373<br>(N=294) | Placebo<br>(N=93) | SII-NVX-CoV2373<br>(N=290) | Placebo<br>(N=96) |
|  |  | Anti-S IgG antibodies |  |  |  |
| 22 days after first dose | N Evaluated | 293 | 93 | 289 | 96 |
|  | n (%) | 293 (100.0) | 5 (5.4) | 281 (97.2) | 4 (4.2) |
|  | 95% CI | (98.7, 100.0) | (1.8, 12.1) | 94.6, 98.8 | 1.1, 10.3 |
| 14 days after second dose | N Evaluated | 293 | 91 | 287 | 96 |
|  | n (%) | 293 (100.0) | 7 (7.7) | 285 (99.3) | 3 (3.1) |
|  | 95% CI | (98.7, 100.0) | (3.1, 15.2) | 97.5, 99.9 | 0.6, 8.9 |
| 180 days after first dose | N Evaluated | 284 | 86 | 282 | 59 |
|  | n (%) | 273 (96.1) | 41 (47.7) | 268 (95.0) | 45 (76.3) |
|  | 95% CI | (93.2, 98.1) | (36.8, 58.7) | (91.8, 97.3) | (63.4, 86.4) |
| Neutralizing antibodies |  |  |  |  |  |
| 22 days after first dose | N Evaluated | 88 | 26 | 131 | 46 |
|  | n (%) | 88 (100.0) | 4 (15.4) | 127 (96.9) | 11 (23.9) |
|  | 95% CI | (95.9, 100.0) | (4.4, 34.9) | (92.4, 99.2) | (12.6, 38.8) |
| 14 days after second dose | N Evaluated | 293 | 91 | 286 | 94 |
|  | n (%) | 291 (99.3) | 10 (11.0) | 282 (98.6) | 7 (7.4) |
|  | 95% CI | (97.6, 99.9) | (5.4, 19.3) | (96.5, 99.6) | (3.0, 14.7) |
| 180 days after first dose | N Evaluated | 85 | 24 | 96 | 17 |
|  | n (%) | 82 (96.5) | 10 (41.7) | 91 (94.8) | 13 (76.5) |
|  | 95% CI | (90.0, 99.3) | (22.1, 63.4) | (88.3, 98.3) | (50.1, 93.2) |

**Table S3:** IgG geometric mean titres after two doses of vaccination for SARS-CoV-2 strains by study day for participants receiving SII-NVX-COV2373.

| Age Group | Parameter | IgG GMTs and GMFR |  |  |  |  |  |
| --- | --- | --- | --- | --- | --- | --- | --- |
|  | Strain | Delta |  |  | Omicron BA.1 |  |  |
|  | Study Day | Baseline | Day 36 | Day 180 | Baseline | Day 36 | Day 180 |
| 2-11 years<br>(n=35) | **GMT**<br>**(95% CI)** | 2433.7<br>(1424.5, 4158.0) | 251510.2<br>(204159.5, 309843.0) | 38347.0<br>(27424.8, 53619.1) | 1080.3<br>(704.3, 1656.9) | 105031.3<br>(83738.1, 131739.0) | 22536.7<br>(17587.9, 28878.0) |
|  | **GMFR**<br>**(95% CI)** | - | 103.3<br>(61.6, 173.4) | 16.9<br>(9.5, 30.2) | - | 97.2<br>(67.1, 140.8) | 22.2<br>(14.9, 33.0) |
| 12-17 years<br>(n=36) | **GMT**<br>**(95% CI)** | 2158.8<br>(1203.4, 3872.7) | 157379.2<br>(122688.8, 201878.3) | 55795.3<br>(41836.2, 74411.9) | 860.2<br>(563.9, 1312.2) | 74971.0<br>(57583.6, 97608.5) | 32008.0<br>(23761.3, 43116.9) |
|  | **GMFR**<br>**(95% CI)** | - | 72.9<br>(41.9, 127.0) | 25.8<br>(13.4, 49.9) | - | 87.2<br>(57.3, 132.6) | 37.2<br>(21.7, 63.7) |

| Age Group | Parameter | IgG GMTs and GMFR |  |  |
| --- | --- | --- | --- | --- |
|  | Strain | Omicron BA.5 |  |  |
|  | Study Day | Baseline | Day 36 | Day 180 |
| 2-11 years<br>(n=35) | **GMT**<br>**(95% CI)** | 1055.3<br>(677.8, 1642.9) | 97840.4<br>(80494.6, 118923.9) | 23087.7<br>(17941.7, 29709.7) |
|  | **GMFR**<br>**(95% CI)** |  | 92.7<br>(62.2, 138.1) | 23.2<br>(14.5, 37.1) |
| 12-17 years<br>(n=36) | **GMT**<br>**(95% CI)** | 793.3<br>(506.7, 1242.0) | 71869.4<br>(56343.4, 91673.9) | 32456.0<br>(23936.3, 44008.0) |
|  | **GMFR**<br>**(95% CI)** |  | 90.6<br>(59.2, 138.6) | 40.9<br>(22.7, 73.9) |

**Table S4:** hACE2 receptor binding inhibition geometric mean titres after two doses of vaccination for Wuhan and variant SARS-CoV-2 strains by study day for participants receiving SII-NVX-COV2373.

| Age Group | Parameter | hACE2 Inhibition titre of antibodies |  |  |  |  |  |
| --- | --- | --- | --- | --- | --- | --- | --- |
|  | Strain | Wuhan |  |  | Delta |  |  |
|  | Study Day | Baseline | Day 36 | Day 180 | Baseline | Day 36 | Day 180 |
| 2-11 years<br>(n=35) | GMT<br>(95% CI) | 10.5<br>(7.8, 14.2) | 704.6<br>(581.5, 853.9) | 129.9<br>(95.8, 176.0) | 12.6<br>(9.1, 17.6) | 538.9<br>(437.3, 664.2) | 131.7<br>(99.0, 175.2) |
|  | GMFR<br>(95% CI) | - | 67.0<br>(49.3, 91.1) | 13.2<br>(9.1, 19.0) | - | 42.7<br>(31.2, 58.5) | 11.1<br>(7.6, 16.1) |
| 12-17 years<br>(n=36) | GMT<br>(95% CI) | 9.5<br>(7.1, 12.7) | 464.5<br>(363.8, 593.1) | 169.4<br>(130.4, 220.1) | 11.6<br>(8.6, 15.7) | 337.0<br>(262.7, 432.3) | 211.7<br>(161.8, 276.9) |
|  | GMFR<br>(95% CI) | - | 48.9<br>(33.9, 70.6) | 17.8<br>(12.2, 26.1) | - | 29.0<br>(20.3, 41.5) | 18.2<br>(12.6, 26.3) |
|  | Strain | Omicron BA.1 |  |  | Omicron BA.5 |  |  |
|  | Study Day | Baseline | Day 36 | Day 180 | Baseline | Day 36 | Day 180 |
| 2-11 years<br>(n=36) | GMT<br>(95% CI) | 6.6<br>(5.4, 8.2) | 173.7<br>(122.0, 247.2) | 57.3<br>(41.8, 78.5) | 7.9<br>(6.1, 10.2) | 229.4<br>(163.7, 321.5) | 91.9<br>(68.6, 123.0) |
|  | GMFR<br>(95% CI) | - | 26.2<br>(18.3, 37.6) | 9.0<br>(6.4, 12.6) | - | 29.1<br>(21.1, 40.3) | 12.1<br>(8.6, 17.0) |
| 12-17 years<br>(n=36) | GMT<br>(95% CI) | 6.0<br>(5.0, 7.2) | 122.4<br>(86.9, 172.4) | 77.1<br>(56.3, 105.6) | 6.5<br>(5.3, 8.0) | 158.1<br>(108.9, 229.6) | 137.6<br>(101.0, 187.5) |
|  | GMFR<br>(95% CI) | - | 20.4<br>(14.2, 29.4) | 12.8<br>(9.0, 18.4) | - | 24.2<br>(16.6, 35.5) | 21.1<br>(14.4, 30.8) |

**Table S5:**
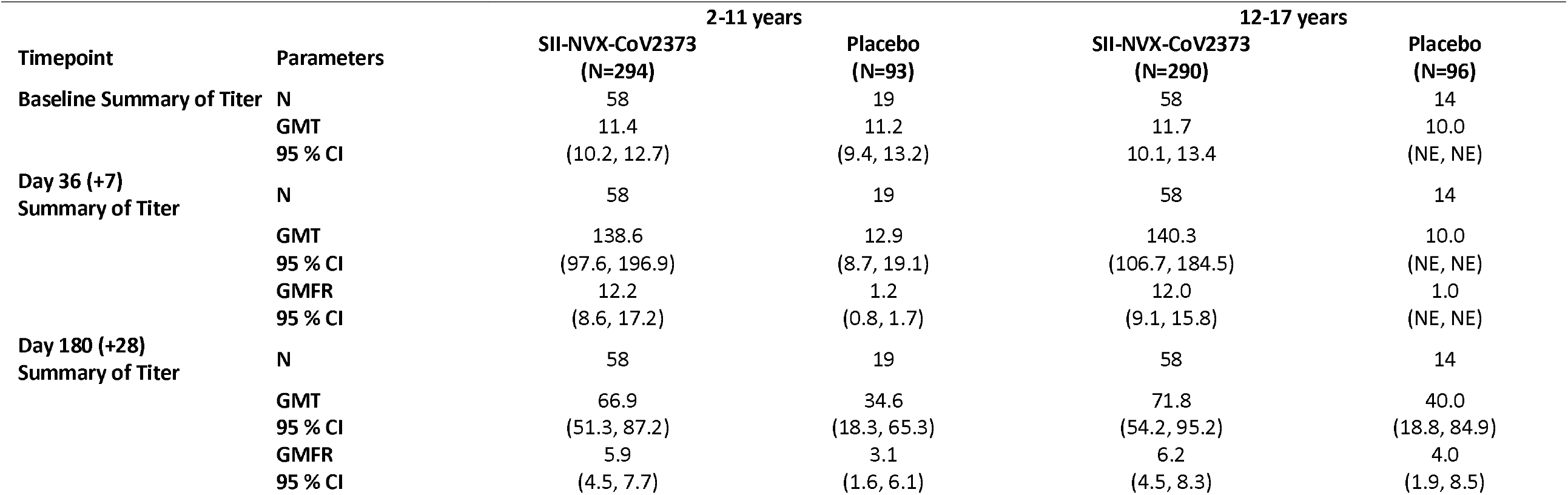
Summary of neutralizing antibodies against SARS-CoV-2 Omicron B.1.1.529 lineage (BA.1) (Immunogenicity Analysis Population)

|  |  |  |  |  |  |
| --- | --- | --- | --- | --- | --- |
| Timepoint | Parameters | 2-11 years |  | 12-17 years |  |
|  |  | SII-NVX-CoV2373<br>(N=294) | Placebo<br>(N=93) | SII-NVX-CoV2373<br>(N=290) | Placebo<br>(N=96) |
| Baseline Summary of Titer | N | 58 | 19 | 58 | 14 |
|  | GMT | 11.4 | 11.2 | 11.7 | 10.0 |
|  | 95 % CI | (10.2, 12.7) | (9.4, 13.2) | 10.1, 13.4 | (NE, NE) |
| Day 36 (+7)<br>Summary of Titer | N | 58 | 19 | 58 | 14 |
|  | GMT | 138.6 | 12.9 | 140.3 | 10.0 |
|  | 95 % CI | (97.6, 196.9) | (8.7, 19.1) | (106.7, 184.5) | (NE, NE) |
|  | GMFR | 12.2 | 1.2 | 12.0 | 1.0 |
|  | 95 % CI | (8.6, 17.2) | (0.8, 1.7) | (9.1, 15.8) | (NE, NE) |
| Day 180 (+28)<br>Summary of Titer | N | 58 | 19 | 58 | 14 |
|  | GMT | 66.9 | 34.6 | 71.8 | 40.0 |
|  | 95 % CI | (51.3, 87.2) | (18.3, 65.3) | (54.2, 95.2) | (18.8, 84.9) |
|  | GMFR | 5.9 | 3.1 | 6.2 | 4.0 |
|  | 95 % CI | (4.5, 7.7) | (1.6, 6.1) | (4.5, 8.3) | (1.9, 8.5) |

